# MedDRA Adoption and Adverse Event Reporting Quality in Gastrointestinal and Abdominal Surgery Randomized Controlled Trials: A Cross-Sectional Analysis

**DOI:** 10.64898/2026.02.04.26345608

**Authors:** Nicholas Camasso, Kohl Kirby, Noah Calvert, John Stroup, Ryan Langerman, Matt Vassar

**Affiliations:** Office of Medical Student Research, Oklahoma State University Center for Health Sciences, Tulsa, Oklahoma, United States; Department of Psychiatry and Behavioral Sciences, Oklahoma State University Center for Health Sciences, Tulsa, Oklahoma, United States

**Keywords:** General Surgery, Harm Minimization, Clinical Practice Guideline, Registry Data

## Abstract

**Introduction:** Adverse event (AE) reporting transparency is essential for evidence-based surgical practice, yet substantial reporting gaps persist despite Consolidated Standards for Reporting Trials (CONSORT) Harms guidance. The Medical Dictionary for Regulatory Activities (MedDRA) provides standardized terminology for AE classification, but its association with AE reporting quality remains unexplored.

**Objectives:** The purpose of this study was to establish the frequency of Medical Dictionary for Regulatory Activities (MedDRA) utilization in gastrointestinal and abdominal surgical trials, identify predictors of its adoption, and quantify the association between MedDRA use and adverse event reporting quality as measured by Completeness scores, registry-publication Concordance, and overall Transparency indices.

**Design:** Cross sectional analysis of matched randomized controlled trial registry-publication pairs.

**Participants:** 116 gastrointestinal and abdominal surgery randomized controlled trials registered on ClinicalTrials.gov with results posted between September 2009 and December 2024 and an associated peer-reviewed publication.

**Primary and Secondary Outcome Measures:** Primary outcomes were differences in AE reporting quality between MedDRA-documenting and non-documenting trials, measured using Harms Reporting Completeness score (0-8), Concordance score (0-7), and Harms Transparency Index (0-15). Secondary outcomes included prevalence of MedDRA adoption and predictors of MedDRA documentation via univariable logistic regression.

**Results:** Among 116 included trials, only 22 (18.8%) explicitly documented MedDRA use. Industry-funded trials (OR=29.32, 95% CI=8.94-118.50, p<0.001) and those with at least one U.S. site (OR=4.59, 95% CI=1.22-30.02, p=0.050) demonstrated significantly higher rates of MedDRA adoption. Trials documenting MedDRA use demonstrated significantly improved reporting across all three score parameters: Completeness score (p<0.001), Concordance score (p=0.002), and Transparency Index (p<0.001). MedDRA use was also associated with lower rates of registry-publication discordance across key safety metrics: serious adverse event (SAE) participant count registry-publication discordance was 59.1% in MedDRA documenting trials and 85.1% in non-MedDRA trials; mortality reporting discordance was 60.0% in MedDRA trials and 82.1% in non-MedDRA trials.

**Conclusion:** Despite strong association with improved AE reporting completeness and registry-publication concordance, MedDRA adoption in gastrointestinal and abdominal surgical trials remains below 20%, concentrated among industry-funded studies. The predominance of unstandardized terminology and free-text strategies promotes reporting inadequacies that complicate evidence synthesis and undermine evidence-based surgical practice. Journals, funding agencies, academic institutions, and researchers should prioritize the adoption of standardized AE terminology to enhance transparency and improve surgical research.

**Trial Registration:** PROSPERO CRD420251081191.

**Strengths and Limitations of this Study:**

- This is the first study to quantify the association between MedDRA use and adverse event reporting quality in surgical trials
- Dual independent screening and extraction with pre-registered protocol minimizes bias and enhances reproducibility
- Analysis limited to gastrointestinal and abdominal surgery; generalizability to other surgical subspecialties remains uncertain
- Required explicit MedDRA documentation; trials using MedDRA without disclosure would be misclassified as non-users
- Concordance assessment examined numerical agreement without evaluating clinical significance of discrepancies

## INTRODUCTION

Randomized controlled trials (RCTs) form the foundation of evidence-based surgical practice, and inform clinical practice guidelines (CPG) such as those published by the American College of Surgeons, European Association for the Study of the Liver (EASL) and American Society of Colon and Rectal Surgeons. [1–3] However, RCTs are only as valuable as their adverse event (AE) reporting. Despite CONSORT harms guidance,[4] substantial heterogeneity persists across surgical and medical trials; whether evaluating procedures, nerve blocks, or perioperative strategies.[5]

Discordance between ClinicalTrials.gov registries and their associated peer-reviewed publication has been demonstrated across a variety of fields,[6–15] representing a critical concern within the broader field of medical research. If physicians are basing standards of care on published research, such research must provide transparent adverse event reporting. Furthermore, the terminology used to report the same AEs differs between, and sometimes even within, studies,[16] complicating the interpretation of AE information. Such discrepancies may undermine confidence in results and alter meta-analyses, ultimately affecting surgical decision making and patient outcomes. The Medical Dictionary for Regulatory Activities (MedDRA), is a standardized and hierarchical terminology for AE classification.[17] Previously, there has been scant investigation into its efficacy as a tool for limiting AE reporting discrepancies in surgical RCTs.

This study aimed to establish the frequency of MedDRA utilization across gastrointestinal and abdominal RCTs and evaluate the link between its use and AE reporting completeness. By analyzing matched registry-publication pairs, we sought to identify factors associated with standardized terminology adoption and quantify its impact on surgical research reporting practices.

## METHODS

### Study Framework and Protocol Registration

This investigation constitutes a planned secondary analysis of a larger cross-specialty project evaluating AE reporting in clinical trials. The overarching study received PROSPERO pre-registration (ID:CRD420251081191).[18] Comprehensive protocol documentation, including predetermined condition classifications, was uploaded to the Open Science Framework (OSF) before data extraction began.[19] The current analysis targets trials related to gastrointestinal and abdominal surgery. Under 45 CFR 46.102(d) and (f), this work did not meet criteria for human subjects research and required no institutional review board approval.[20]

No patients or members of the public were involved in the design, conduct, reporting, or dissemination of this research. This study constitutes a meta-research analysis of publicly available trial registry and publication data, examining adverse event reporting practices rather than clinical outcomes or patient-centered research questions.

### Search Methodology

On June 28, 2025, the parent investigation executed a systematic ClinicalTrials.gov search, capturing interventional Phase 2–4 and “Not Applicable” trials with results posted from September 27, 2009, through December 31, 2024, consistent with FDAAA Final Rule timelines.[21] We used a manually constructed Boolean search strategy containing surgical terms, identified synonyms, and conceptually related terms to avoid the registry’s automated term expansion functionality. Associated publications were located via ClinicalTrials.gov linkage, supplemented by manual PubMed and Google Scholar searches utilizing registry NCT, title, and other identifiers, for registry-associated manuscripts. This process was completed by two independent researchers and documented in Google Sheets. Obtained articles were deduplicated after article comparison. Other registries, such as the International Clinical Trials Registry Platform (ICTRP), and unpublished literature were not consulted. ClinicalTrials.gov served as the exclusive registry given its unique structure, and numerically defined AE fields enabling cross-platform comparison.

### Inclusion and Exclusion Standards

We included trials registered on ClinicalTrials.gov that evaluated at least one primary intervention or perioperative strategy related to gastrointestinal or abdominal surgery, had publicly posted results, and an associated peer-reviewed publication. Eligible interventions encompassed procedural approaches (e.g., bariatric surgery, colectomy, cholecystectomy, hernia repair) or perioperative strategies (e.g., analgesic techniques, surgical devices, fluid management protocols). Trials were excluded if they lacked a corresponding peer-reviewed publication, were Phase 1 or observational studies, reported only a trial protocol, were published in a non-English language, lacked surgical AE data, or focused on non-gastrointestinal or abdominal surgery. The PRISMA diagram of the parent investigation is publicly accessible on OSF.[19]

### Trial Selection and Surgery Classification

Two independent reviewers screened all registry entries and publications according to predetermined eligibility standards. Inclusion of studies involved dual independent review with disagreement resolution through consensus discussion or third reviewer arbitration. This secondary analysis required no additional screening or data collection beyond the original investigation.

### Data Collection Protocol

The parent database originated from dual independent extraction utilizing a standardized, pilot-tested Google Form. Extractors completed structured training sessions and calibration exercises before commencing full-scale extraction. Variables captured encompassed trial features such as: sponsorship, intervention classification, participant numbers, and trial phase as well as comprehensive AE-related elements from both ClinicalTrials.gov and matched publications. AE language was documented inclusively, encompassing variant equivalent terminology such as “complications” or “undesirable effects,” consistent with 42 CFR §11.10(a) regulatory definitions.[21] Presentation ambiguities were documented and resolved during extraction. Data for the current analysis were obtained directly from this parent dataset.

### Primary Exposure: MedDRA Documentation

The principal exposure involved explicit citation of MedDRA use within either the ClinicalTrials.gov results or the publication. Direct textual mention was mandatory; presumed or implicit usage was not recognized. Trials were stratified as registry-only, publication-only, both sources, or absent MedDRA documentation from both the registry and publication. For regression modeling, MedDRA documentation was dichotomized as present versus absent.

### Study Outcomes: Harms Reporting Completeness Score

The principal outcome comprised an 8-component Harms Reporting Completeness Score developed from CONSORT Harms guidelines and the extraction framework structure.[22] Publications earned one point for documenting each element: SAE participant counts, SAE event counts, OAE participant counts, OAE event counts, explicit OAE frequency threshold, any AE results format (descriptive or tabular), AE-related treatment discontinuations, and all-cause mortality. Total scores spanned 0 to 8.

### Registry–Publication Agreement: Concordance Score

Agreement between ClinicalTrials.gov and publications was evaluated across the seven AE dimensions: SAE participant counts, SAE event counts, OAE participant counts, OAE event count, AE-related discontinuations, mortality, and OAE threshold (similar to the Harms Reporting Completeness Score minus the “any AE results format”). Dimensions achieved concordance when numerically equivalent across platforms. Trials lacking data in both sources for each specific dimension were omitted from that dimension’s denominator and therefore did not contribute to percentage calculations. Total Concordance Scores ranged 0 to 7.

### Combined Harms Transparency Index

An integrated index combined the Completeness Score (0–8) and Concordance Score (0–7), producing a Harms Transparency Index spanning 0 to 15.

### Predictor Analysis of MedDRA Documentation

Univariable logistic regression examined predictors of MedDRA documentation (yes vs no). Predictor variables, selected prior to data extraction, included trial phase (e.g. Phase 2), sponsorship (e.g. non-industry), AE burden (high vs low, determined by median total AE count from ClinicalTrials.gov), log10-transformed enrollment, and U.S. site involvement. Complete case analysis proceeded without imputation.

### Analytical Approach

Descriptive statistics characterized trial features and outcome distributions. Wilcoxon rank-sum tests assessed differences in Completeness, Concordance, and Harms Transparency Index scores between MedDRA-documenting and non-documenting trials. Concordance percentages reflected only evaluable trials. All analyses utilized R (version 4.3 or later).

### Bias Mitigation and Protocol Adherence

Since the goal centered on evaluating documentation practices rather than clinical endpoints, conventional risk-of-bias instruments were not employed. Bias mitigation strategies included duplicate screening, duplicate extraction, reviewer training, and predetermined analytic methods. Parent protocol deviations are documented on OSF; no deviations occurred for this surgery MedDRA analysis.[19]

### Transparency and Resource Availability

All data, code, scoring rubrics, complete protocol documentation, and the PRISMA diagram of the parent study are publicly accessible through OSF.[19] This investigation follows best practices for transparency, reproducibility, and open meta-research standards.

## RESULTS

### General Characteristics of Included Trials

Supplementary Table 1 describes the general characteristics of included trials, including trial phase, allocation and enrollment, location, funding source, intervention type, MedDRA use and AE reporting frequency. Notably, of the 116 included studies, 18 (15.5%) reported the use of MedDRA solely in ClinicalTrials.gov, 1 (0.9%) reported the use of MedDRA strictly in the publication, 3 (2.6%) studies reported MedDRA use in both the registry and publication, and 94 (81%) did not report MedDRA use in either data source.

### Registry-Publication Concordance in AE Reporting

Registry-publication concordance and discordance in AE reporting across the seven AE dimensions compromising the concordance score (SAE participant counts, SAE event counts, OAE participant counts, OAE event count, AE-related discontinuations, mortality, and OAE threshold) is available in Table 1. Most commonly, discordance was noted for each domain, with only AE-related discontinuations more commonly not reported in either source. The greatest discordance was found in the OAE reporting threshold, with 111 (94.9%) trials being discordant in this reported value. OAE participant counts also showed high discordance, with 105 (89.7%) studies reporting differing information between their registry and publication. The most commonly reported concordant statistic was SAE participant counts, where 23 (19.7%) trials reported identical information in both sources.

**Table 1.** Registry–Publication Concordance Across Adverse Event Reporting Domains. Data represent the number and percentage of trials (N=116) demonstrating concordance (identical information reported in both registry and publication), discordance (differing information between sources), or absence of reporting in both sources across seven adverse event dimensions. The OAE reporting threshold refers to the percentage cut-off used to determine which adverse events were reported in detail.

| Domain | Concordant trials (%) | Discordant trials (%) | Neither source reported (%) |
| --- | --- | --- | --- |
| SAE participant counts | 23 (19.8) | 93 (80.2) | 0 (0.0) |
| SAE event counts | 9 (7.8) | 64 (55.2) | 43 (37.1) |
| Mortality (all-cause) | 20 (17.2) | 67 (57.8) | 29 (25.0) |
| OAE participant counts | 11 (9.5) | 105 (90.5) | 0 (0.0) |

| **Table 1. Registry–Publication Concordance Across Adverse Event Reporting Domains** |  |  |  |
| --- | --- | --- | --- |
| OAE event counts | 7 (6.0) | 73 (62.9) | 36 (31.0) |
| AE-related discontinuations | 2 (1.7) | 30 (25.9) | 84 (72.4) |
| OAE reporting threshold | 5 (4.3) | 111 (95.7) | 0 (0.0) |

### Registry-Publication Concordance in AE and Mortality Reporting by MedDRA Use

Discordance in SAE participants, SAE events, and mortality is available in Table 2A. Of the 116 total included trials, only 22 reported any MedDRA use in either CT.gov or the publication. Of those twenty-two trials, 13 (59.1%) were discordant in SAE participant counts, while 80 (85.1%) of the 94 trials that did not report MedDRA use were discordant in SAE participants. These two groups also differed in percent of reported SAE event counts: 17 (81%) trials with any MedDRA use and 47 (90.4%) trials with no MedDRA use were discordant in this category. MedDRA use was also associated with lower mortality reporting discordance (60.0% vs. 82.1%).

Discordance in the other domains (i.e. OAE participants, OAE events, Treatment discontinuation, and OAE threshold) is available in Table 2B. Among trials reporting MedDRA use, discordance between registry and publication occurred in 90.9% for OAE participants, 84.2% for OAE events, 81.8% for treatment discontinuation, and 95.5% for OAE threshold. When all trials are considered, regardless of MedDRA reporting status, mortality discordance was lowest with 67 (77.0%) trials.

A visual comparison of discordance across all domains is present in Figure 1.

**Table 2A.** Registry–Publication Discordance in Serious Adverse Event and Mortality Reporting by MedDRA Use. Data represent the number and percentage of trials demonstrating discordance (differing information between registry and publication) across three reporting domains, stratified by whether trials reported the use of MedDRA terminology in the registry (CT.gov), publication, both sources, or neither source. Percentages are calculated among trials where at least one source reported data for the respective domain.

| **Table 2A. Registry–Publication Discordance in Serious Adverse Event and Mortality Reporting by MedDRA Use** |  |  |  |  |
| --- | --- | --- | --- | --- |
| MedDRA use group | N | SAE participants discordant, n (%) | SAE events discordant, n (%) | Mortality discordant, n (%) |
| CT.gov only | 18 | 11 (61.1) | 14 (77.8) | 10 (62.5) |
| Publication only | 1 | 1 (100.0) | 1 (100.0) | 1 (100.0) |
| Both | 3 | 1 (33.3) | 2 (100.0) | 1 (33.3) |
| Any MedDRA use | 22 | 13 (59.1) | 17 (81.0) | 12 (60.0) |
| No MedDRA use | 94 | 80 (85.1) | 47 (90.4) | 55 (82.1) |
| Overall | 116 | 93 (80.2) | 64 (87.7) | 67 (77.0) |

Note: Percentages exclude trials not reporting the outcome in either source.

**Table 2B.** Registry–Publication Discordance in Other Adverse Event Domains by MedDRA Use. Data represent the number and percentage of trials demonstrating discordance (differing information between registry and publication) across four reporting domains for other adverse events, stratified by whether trials reported the use of MedDRA terminology in the registry (CT.gov), publication, both sources, or neither source. Percentages are calculated among trials where at least one source reported data for the respective domain.

| **Table 2B. Registry–Publication Discordance in Other Adverse Event Domains by MedDRA Use** |  |  |  |  |  |
| --- | --- | --- | --- | --- | --- |
| MedDRA use group | N | OAE participants discordant, n (%) | OAE events discordant, n (%) | Treatment discontinuation discordant, n (%) | OAE threshold discordant, n (%) |
| CT.gov only | 18 | 16 (88.9) | 13 (81.2) | 8 (80.0) | 17 (94.4) |
| Publication only | 1 | 1 (100.0) | 1 (100.0) | 0 (0.0) | 1 (100.0) |
| Both | 3 | 3 (100.0) | 2 (100.0) | 1 (100.0) | 3 (100.0) |
| Any MedDRA use | 22 | 20 (90.9) | 16 (84.2) | 9 (81.8) | 21 (95.5) |
| No MedDRA use | 94 | 85 (90.4) | 57 (93.4) | 21 (100.0) | 90 (95.7) |
| Overall | 116 | 105 (90.5) | 73 (91.2) | 30 (93.8) | 111 (95.7) |

Note: Percentages exclude trials not reporting the outcome in either source.

**Figure 1.**
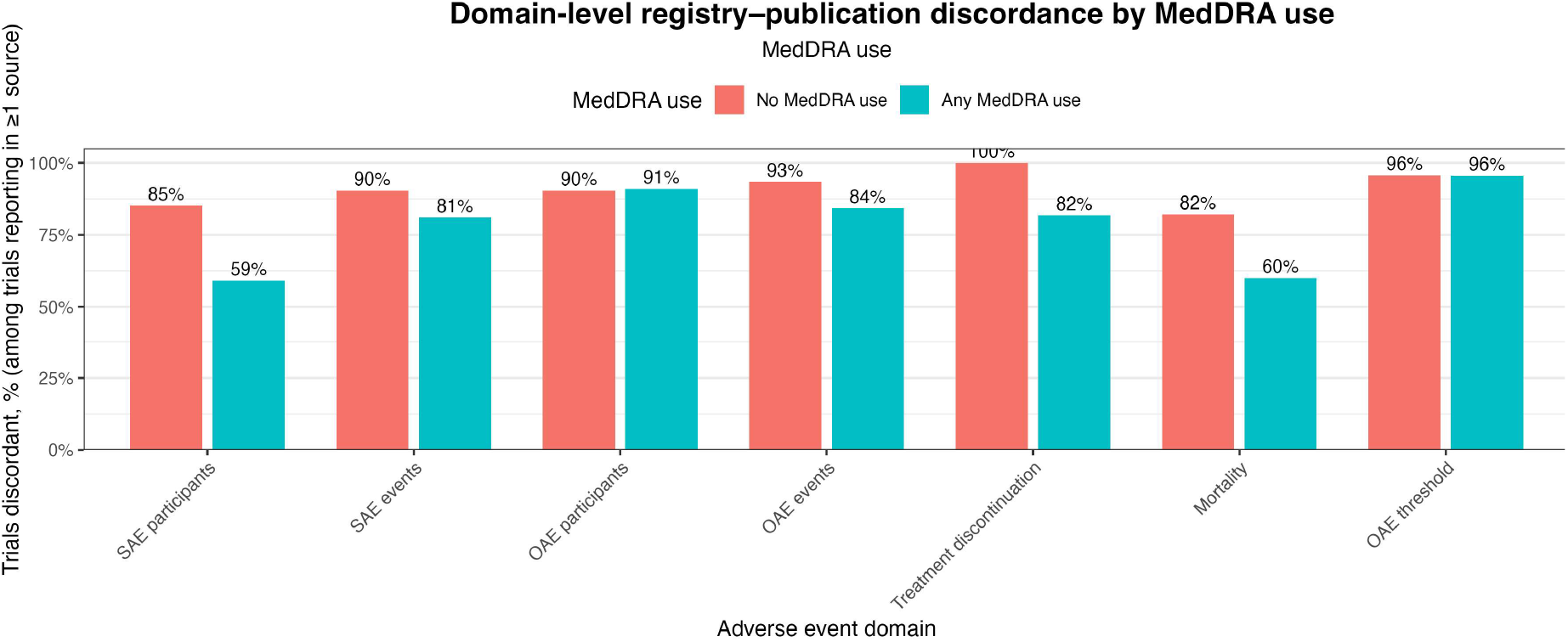
Domain-level registry–publication discordance by MedDRA use. Bar chart comparing the percentage of trials with discordant adverse event reporting (differing information between registry and publication) across the seven reporting domains, stratified by reported use of MedDRA terminology. Red bars represent trials with no MedDRA use in either source; teal bars represent trials with any reported MedDRA use.

### Completeness, Concordance, and Transparency Index Score

Median combined Harms Transparency Index scores, ranging from 0-15, and their IQR for each MedDRA group (e.g. CT.gov only, Publication only, etc) as well as p-values for comparison between groups are reported in Table 3A. Completeness scores were highest in the CT.gov only, 5.0 (IQR 3.2-7.0) and the any MedDRA use, 5.0 (IQR 4.0-7.0) categories. Those that did not report any MedDRA use had a median Completeness score of 2.0 (IQR 0.0-5.0). Concordance scores were highest in the group of three trials that reported MedDRA use in both the registry and publication; these trials had a median score of 2.0 (IQR 1.0-2.0). The Transparency index was greatest in the “any MedDRA use” trial group, 7.0 (IQR 4.2-7.0). Trials without any reported MedDRA use had a median transparency index of 3.5 (IQR 1.0-6.0).

**Table 3A.**
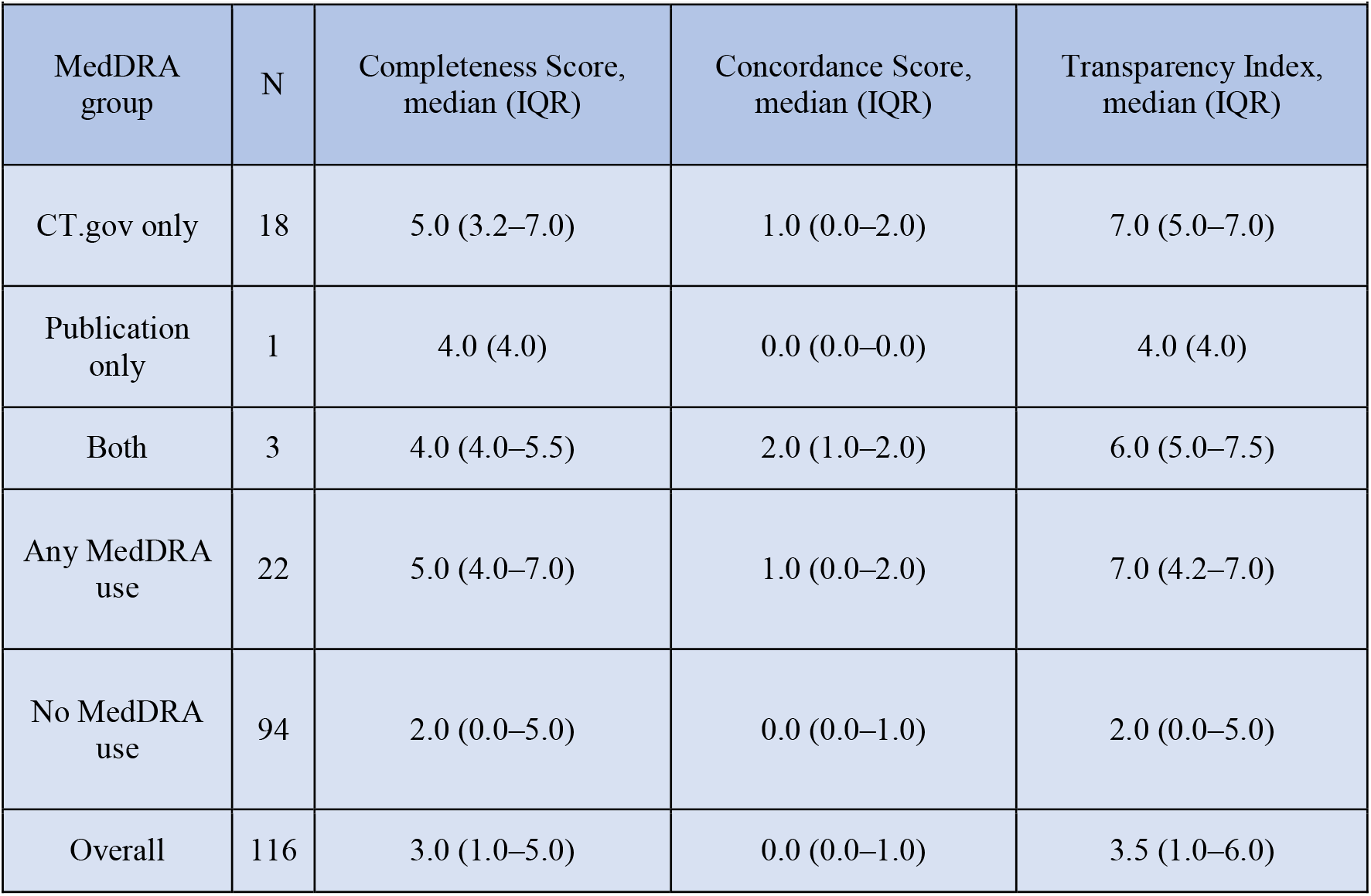
Completeness, Concordance, and Transparency Index by MedDRA Use. Data represent median scores with interquartile ranges (IQR) for three metrics assessing adverse event reporting quality, stratified by whether the use of MedDRA terminology was reported in the registry (CT.gov), publication, both sources, or neither source. The Completeness Score (range 0–14) reflects the total number of adverse event domains reported across both sources. The Concordance Score (range 0–7) reflects the number of domains with identical information in both sources. The Transparency Index (range 0–14) sums the Completeness and Concordance scores.

| **Table 3A. Completeness, Concordance, and Transparency Index by MedDRA Use** |  |  |  |  |
| --- | --- | --- | --- | --- |
| MedDRA group | N | Completeness Score, median (IQR) | Concordance Score, median (IQR) | Transparency Index, median (IQR) |
| CT.gov only | 18 | 5.0 (3.2–7.0) | 1.0 (0.0–2.0) | 7.0 (5.0–7.0) |
| Publication only | 1 | 4.0 (4.0) | 0.0 (0.0–0.0) | 4.0 (4.0) |
| Both | 3 | 4.0 (4.0–5.5) | 2.0 (1.0–2.0) | 6.0 (5.0–7.5) |
| Any MedDRA use | 22 | 5.0 (4.0–7.0) | 1.0 (0.0–2.0) | 7.0 (4.2–7.0) |
| No MedDRA use | 94 | 2.0 (0.0–5.0) | 0.0 (0.0–1.0) | 2.0 (0.0–5.0) |
| Overall | 116 | 3.0 (1.0–5.0) | 0.0 (0.0–1.0) | 3.5 (1.0–6.0) |

Wilcoxon rank-sum tests (Table 3B) comparing the distribution of each MedDRA group to the no MedDRA use group, identified significantly higher scores in Completeness, Concordance, and Harms Transparency Index. Comparison of the CT.gov only group with the no MedDRA group displayed significance across completeness scores (p<0.001), concordance scores (p=0.002), and transparency index (p<0.001). Comparison of the any MedDRA use group to the no MedDRA use group showed similar results, with all three sections having significant p-values. A graphical comparison of the no MedDRA use group and any MedDRA use group is presented in Figure 2.

**Table 3B.** Comparison of Score Distributions by MedDRA Use Group. Data represent p-values from Wilcoxon rank-sum tests comparing Completeness Score, Concordance Score, and Transparency Index distributions between trials reporting the use of MedDRA terminology and trials not using MedDRA. Each MedDRA use category (CT.gov only, publication only, both sources, or any MedDRA use) was compared against the reference group of trials with no MedDRA use in either source. P-values <0.05 indicate statistically significant differences in score distributions.

| Table 3B. Comparison of Score Distributions by MedDRA Use Group |  |  |  |
| --- | --- | --- | --- |
| Comparison | Completeness p-value | Concordance p-value | Transparency Index p-value |
| CT.gov only vs No MedDRA use | <0.001 | 0.002 | <0.001 |
| Publication only vs No MedDRA use | 0.540 | 0.540 | 0.670 |
| Both vs No MedDRA use | 0.094 | 0.122 | 0.072 |
| Any MedDRA use vs No MedDRA use | <0.001 | 0.002 | <0.001 |

P-values: Wilcoxon rank-sum tests comparing each MedDRA group with trials that did not use MedDRA.

**Figure 2.**
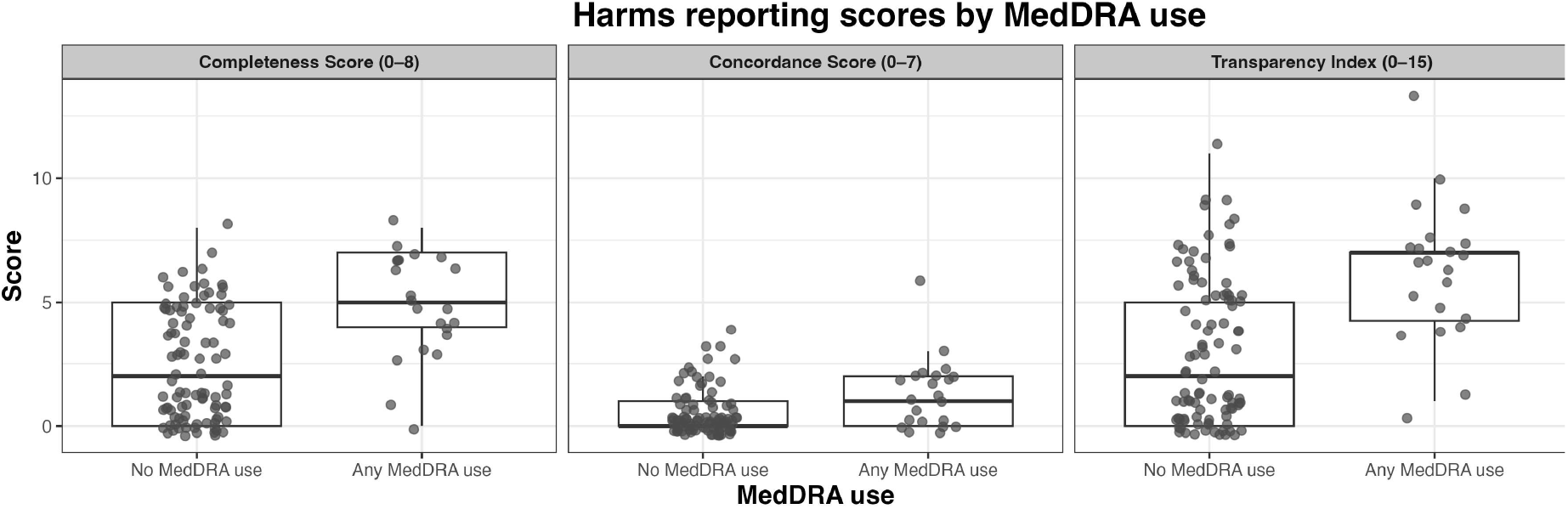
Harms reporting scores by MedDRA use. Individual data points with overlying Box plots comparing adverse event reporting quality metrics between trials with any reported use of MedDRA terminology (in registry, publication, or both) versus trials with no reported MedDRA use in either source. Left panel: Completeness Score (range 0–8) represents the number of adverse event domains reported across both registry and publication. Middle panel: Concordance Score (range 0–7) represents the number of domains with identical information between sources. Right panel: Transparency Index (range 0–15) represents the sum of Completeness and Concordance scores. Box plots show median (center line), interquartile range (box), and range excluding outliers (whiskers). Individual trials are represented by gray dots.

### Univariable predictors of MedDRA use

Univariable predictors of MedDRA, including trial phase, funding source, AE burden, enrollment population, and U.S. site, were examined using univariable logistic regression models (Table 4). Notably, industry-funded trials (OR=29.66, p<0.001), and trials with at least one U.S. site (OR=4.59, p=0.050) had greater odds of utilizing MedDRA classification.

**Table 4.** Univariable Predictors of MedDRA Use. Data represent odds ratios (OR) with 95% confidence intervals (CI) and p-values from univariable logistic regression analyses examining factors associated with the use of MedDRA terminology reported in either the registry or publication. Phase 2 trials, non-industry/other funding, low adverse event burden, and absence of U.S. sites served as reference categories. Enrollment was log-transformed to assess the effect of a 10-fold increase in sample size. P-values <0.05 indicate statistically significant associations.

| Table 4. Univariable Predictors of MedDRA Use |  |  |  |
| --- | --- | --- | --- |
| Predictor Category | Specific Predictor | OR (95% CI) | p-value |
| Trial Phase | Phase 3 vs Phase 2 | 1.22 (0.22–7.40) | 0.818 |
|  | Phase 4 vs Phase 2 | 0.17 (0.01–1.47) | 0.139 |
|  | Not Applicable vs Phase 2 | 0.78 (0.20–3.84) | 0.728 |
| Funding Source | Industry-including vs Non-industry/other | 29.32 (8.94–118.50) | <0.001 |
| AE Burden | High vs Low | 2.72 (0.99–8.30) | 0.061 |

| **Table 4. Univariable Predictors of MedDRA Use** |  |  |  |
| --- | --- | --- | --- |
| **Enrollment** | Log10 enrollment (per 10-fold increase) | 1.59 (0.50–5.16) | 0.429 |
| **U.S. Site** | Yes vs No | 4.59 (1.22–30.02) | 0.050 |

## DISCUSSION

This study’s analysis reveals strikingly low MedDRA adoption in gastrointestinal and abdominal surgery RCTs, with fewer than 20% of trials explicitly documenting its use in either the registry or associated peer-reviewed publication, despite growing calls for transparent and standardized AE reporting. Our findings demonstrate that industry funding and U.S. site involvement are the dominant drivers of MedDRA utilization, with industry funding demonstrating more than 29-times greater likelihood of adoption. Those that did document MedDRA use, however, demonstrated markedly improved adverse event reporting. Completeness scores (5.0 versus 2.0), Concordance scores (1.0 versus 0.0), and Transparency Index scores (7.0 versus 2.0) were all significantly greater, on average, compared to their non-MedDRA-using counterparts. This suggests that the adoption of standardized terminology may be a marker, or possibly a driver, of methodological rigor and AE transparency in surgical research.

These marked differences in AE reporting carry direct implications for clinical practice. AE rates contribute to the development of clinical practice guidelines, inform patient counseling on procedural risks, and contribute to accurate risk-benefit assessments for the practicing surgeon. Regardless of MedDRA association, the magnitude of registry-publication discordance observed in this study contributes to a growing population of literature highlighting AE reporting failures.[23–27] Trials using MedDRA were significantly more likely to report critical elements: serious adverse event counts that distinguish minor perioperative complications from life-threatening issues, explicit frequency thresholds clarifying reporting criteria, and treatment discontinuation rates that signal concern.

MedDRA’s five-level hierarchy offers advantages beyond regulatory compliance. Unlike free-text descriptions, it provides precise event classification, mitigating issues with synonymy and inconsistent categorization. Currently, trialists may classify the same AE under differing high-level categories. For example, wound dehiscence was reported under “injury, poisoning, and procedural complications” in one trial, and “infections and infestations” in another.[28,29] Trials may also vary in their description of the adverse event. While some may take the more descriptive approach, “Non-ST-elevation myocardial infarction “, others provide broader terminology, like “myocardial infarction”.[29,30] Meta-analysts face impossible decisions in determining event equivalence, risking the introduction of subjectivity to the literature, and may therefore be forced to limit analysis to broader classifications by organ system grouping.

Standardized systems, such as MedDRA, limit the inherent ambiguity present in current free-text descriptions, where identical complications are described using different terms and categories across trials. It simplifies meta-analysis, promoting greater statistical summation and more consistent complication data for utilization in evidence-based clinical practice guidelines, patient education, and risk-benefit analysis. It enhances registry-publication concordance by creating classifications that can be reported consistently across platforms without ambiguity. These practices fulfill the ethical obligation to trial participants, physicians, and ultimately to the patients, who rely on reproducible, accessible evidence that advances surgical care.

Industry funding emerged as the dominant predictor of MedDRA adoption, with industry-sponsored trials showing 29-fold greater odds of use compared to non-industry trials. This disparity likely reflects multiple mechanisms: pharmaceutical and device companies face regulatory requirements for standardized AE reporting to agencies like the FDA, possess the financial resources to license MedDRA, and house dedicated data management infrastructure with trained personnel. Academic investigators, conversely, may lack familiarity with MedDRA despite its widespread use within pharmaceutical trials, receive little training in standardized terminology systems, and operate without dedicated safety reporting staff. These findings suggest MedDRA adoption barriers are structural and financial, not intellectual. Mandating use without addressing licensing costs, training infrastructure, and data management support risks creating compliance burdens without enabling implementation.

Addressing the disparity in AE reporting requires coordinated action on the part of researchers, journals, meta-analysts, funding agencies, and clinical practitioners. Journals should mandate standardized AE terminology as a requirement for publication, or prioritize manuscripts documenting standardized terminology as a quality marker analogous to trial registration; funding agencies should require the use of standardized AE terminology as a condition with provisions for MedDRA licensing and training, and academic medical centers should incorporate terminology training into their research fellowship curriculum. While MedDRA utilization alone cannot ensure comprehensive AE reporting, it represents a critical component of transparency infrastructure that researchers, journals, academic medical institutions, and funding agencies should prioritize in surgical research.

### Limitations

Several limitations warrant consideration in interpreting the findings of this study. This analysis focused exclusively on gastrointestinal and abdominal surgery. Its generalizability to other surgical subspecialties and medical RCTs remains uncertain, though widespread reporting challenges, outside of MedDRA use, have been documented previously. This investigation also required explicit MedDRA documentation; trials which utilized this system without outwardly stating its use would be mis-classified as non-users. However, undocumented use itself represents a transparency failure, as readers and editors cannot verify standardized coding without explicit disclosure. Furthermore, our concordance assessment examined numerical agreement without evaluating the clinical impact of these discrepancies; those with 1% discrepancy, and those with 20% discrepancy, were both documented as discordant, though the clinical implication of the discordance is vastly different. MedDRA adoption may also be an indicator of unmeasured factors such as the institutional research culture and data-management infrastructure present at trial sites. We also relied exclusively on ClinicalTrials.gov and published manuscripts, excluding a large population of research that may demonstrate improved AE reporting practices.

## Conclusion

The utilization of MedDRA with its explicit documentation in gastrointestinal and abdominal surgical trials is strongly associated with improved AE reporting completeness, concordance, and overall transparency; yet adoption of MedDRA in this field falls below 20% of trials. Strong indicators of MedDRA use include industry-funded trials and those with at least one U.S. site. As a whole, RCTs in this field report adverse events with insufficient rigor to support evidence-based clinical decision making and reliable evidence synthesis. Current AE reporting inconsistencies undermine patient counseling, guideline development, and evidence-based care, problems that standardized terminology could significantly address. If journals prioritize MedDRA utilization, it encourages funding agencies to mandate and budget for its implementation and prompts academic institutions to incorporate adverse events reporting training into their research curriculum. Without standardized AE reporting, we risk building the future of evidence-based surgery on a foundation of incomparable and unreliable data.

## Data Availability

All data produced in the present work are available online at the Open Science Framework.

https://osf.io/uqpmc/files/osfstorage?view_only=438362990977431989f5f74479965c81

## Contributors

Nicholas Camasso contributed to data interpretation and writing – original draft preparation. Ryan Langerman and Kohl Kirby were responsible for methodology, including drafting the protocol, interpreting results, providing project administration, and conducting formal data analysis. John Stroup and Noah Calvert were responsible for investigation, data curation, and writing – reviewing and editing. Matt Vassar led the conceptualization, supervision, and project administration, provided student training, and contributed to writing – review & editing. Claude (Anthropic) was used to refine sentence structure and word choice during the drafting of the manuscript. Authors provided original text to the AI for editing suggestions. These suggestions were then reviewed and modified as deemed necessary before being implemented in the manuscript. AI was not used for data analysis, to interpret results or to generate scientific conclusions. The authors take full responsibility for this work.

