## Supplemental Table 1 for "MedDRA Adoption and Adverse Event Reporting Quality in Gastrointestinal and Abdominal Surgery Randomized Controlled Trials: A Cross-Sectional Analysis"

**Supplemental Table 1. General Characteristics of Included Trials**

| Characteristic | n (%) |
|---|---|
| **Trial Phase (N=116)** | |
| Not Applicable | 63 (54.3) |
| 4 | 23 (19.8) |
| 3 | 16 (13.8) |
| 2 | 14 (12.1) |
| **Allocation & Enrollment** | |
| Randomized | 96 (82.8) |
| Unspecified | 20 (17.2) |
| Enrollment, median (IQR) | 103 (53–178) |
| **Study Location** | |
| North America | 78 (67.2) |
| Europe | 18 (15.5) |
| Asia | 9 (7.8) |
| Multiple continents | 8 (6.9) |
| Africa | 1 (0.9) |
| Australia/Oceania | 1 (0.9) |
| South America | 1 (0.9) |
| **Funding Source** | |
| Institutional | 85 (73.3) |
| Industry | 26 (22.4) |
| Government | 3 (2.6) |
| Private | 2 (1.7) |
| **Intervention type** | |
| Drug | 51 (44.0) |
| Device | 38 (32.8) |
| Procedure | 19 (16.4) |
| Other | 4 (3.4) |

| Characteristic | n (%) |
|---|---|
| Biological | 2 (1.7) |
| Behavioral | 1 (0.9) |
| Diagnostic_test | 1 (0.9) |
| **MedDRA use** | |
| CT.gov only | 18 (15.5) |
| Publication only | 1 (0.9) |
| Both | 3 (2.6) |
| Neither | 94 (81.0) |
| **AE reporting frequency** | |
| Trials with any SAE | 71 (61.2) |
| Trials with any OAE | 80 (69.0) |
| Trials with any treatment discontinuation due to AEs | 25 (21.6) |
| Trials with any mortality event | 30 (25.9) |
